# Financial incentives to motivate treatment for hepatitis C with direct acting antivirals among Australian adults (The Methodical evaluation and Optimisation of Targeted IncentiVes for Accessing Treatment of Early-stage hepatitis C: MOTIVATE-C): Statistical analysis plan

**DOI:** 10.1101/2024.11.27.24318114

**Authors:** Mark Jones, James Totterdell, Parveen Fathima, Thomas L Snelling

**Affiliations:** Sydney School of Public Health, Faculty of Medicine and Health, The University of Sydney, Sydney, 2050; Wesfarmers Centre of Vaccines and Infectious Diseases, Telethon Kids Institute, Nedlands, 6009

**Keywords:** Bayesian design, Adaptive trial, Hepatitis C, HCV, Dose–response, Financial incentives, Direct-acting antiviral, Randomised study

## Abstract

The MOTIVATE-C study explores a critical question in hepatitis C treatment: How do financial incentives influence patients’ decisions to initiate direct-acting antiviral (DAA) therapy? Using an innovative Bayesian adaptive design, the research aims to determine the precise relationship between monetary support and treatment initiation among individuals with untreated hepatitis C virus. The study’s unique approach involves response-adaptive randomization, which dynamically allocates participants to different financial incentive levels. As the trial progresses, doses more likely to encourage treatment will receive increased emphasis, while less effective incentive levels may be systematically eliminated through pre-defined futility stopping rules. Participants will be tracked for DAA therapy initiation within 12 weeks of enrollment, with dedicated study navigators assisting them through the treatment access process. The primary analysis will adhere to the intention-to-treat principle, ensuring a comprehensive and unbiased evaluation of the intervention’s effectiveness. This manuscript details the statistical analysis plan, presenting the precise methodological framework, decision-making criteria, and analytical thresholds that will guide the study’s interpretation of how financial incentives might overcome barriers to hepatitis C treatment.

## 1 Introduction

The purpose of this document is to describe the statistical approach that will be used in the analysis of data and also the pre-specified adaptations for the Motivate-C study. It is targeted at a statistical audience and should be read in conjunction with the study protocol (Fathima et al. 2024). Any significant variations from what is documented here will be reported with the results.

### 1.1 Background

Infection with hepatitis C virus (HCV) causes acute or chronic hepatitis. While acute HCV infections are usually asymptomatic and can clear spontaneously, chronic HCV can lead to liver damage and death. Healthcare costs associated with chronic HCV infection are high and the condition has been recognised as a major public health issue in Australia. In 2020, approximately 120k people were estimated to be living with chronic HCV. Many of them are believed to be from marginalised societal groups that are under-served by the healthcare system.

Direct-acting antivirals (DAA) are a relatively new and highly effective treatment for all types of hepatitis C that essentially cure patients of the virus. However, while DAA’s have minimal side effects, necessitate only a short time on treatment (8-12 weeks) and have been heavily subsidised by the government in Australia, their uptake remains low.

Financial incentives may be both a viable and cost-effective option to increase the uptake of DAA therapy. The Motivate-C study aims to evaluate the use of direct payment financial incentives to increase the probability of DAA treatment initiation in people with chronic HCV.

### 1.2 Study Objectives

The objectives of this project are to assess, within the context of a patient-support program:

1. Does offering a financial incentive to people with HCV to initiate DAA therapy in primary care, increase the probability of treatment initiation compared to usual care (no incentive)?
2. What is the dose-response relationship for the range of financial incentives offered and the probability of treatment initiation?
3. What participant incentive is optimal in terms of minimising the costs expended under the incentive program relative to the costs averted from the progression of untreated chronic HCV?

Secondary aims of Motivate-C examine the extent to which primary care provider co-incentives modify the probability of treatment initiation. However, the prescriber effects can only be evaluated when the patient nominates a primary care provider, and this means that there may be relatively little data available to estimate them. Specific research questions are:

1. Does offering primary care providers a co-incentive payment modify the probability of treatment initiation?
2. How is the dose-response relationship modified by a co-incentive offered to the primary care provider?
3. What combination of participant and primary care provider incentive is optimal in terms of minimising the costs expended under the incentive program relative to the costs averted from the progression of untreated chronic HCV?

### 1.3 Study Design

Dose-response studies evaluate the relationship between a dose and a subsequent response (Laird et al. 2020). In Motivate-C, the dose corresponds to the magnitude of the financial incentive and the response corresponds to the probability of treatment initiation with DAA. While dose-response studies are usually associated with the pharmacological literature (e.g. Grieve and Krams (2005)), the designs have also been applied in psychology wherein the dose component might refer to the number of therapy sessions delivered or a similarly modifiable intervention (Howard et al. 1986).

A dose-response model could be useful for evaluating the effects of financial incentives. Usually, studies involving financial incentives examine a small number of different monetary reward values. However, this approach presumes that the amounts selected are sufficient to create a detectable response relative to the reference/control state. In such a design, the absence of a treatment effect may erroneously lead to the conclusion that financial incentives systematically do not work when all that was required was a slightly higher incentive. A dose-response design allows us to leverage the correlation in response across a wide range of incentives under an assumed model and will give a way in which to estimate the minimal incentive value that is necessary to induce a minimally important response relative to no incentive.

In Motivate-C, participants will be randomised to one of several pre-specified incentive values with the goal of encouraging DAA therapy initiation in primary care. The Bayesian adaptive dose-ranging design uses a group sequential approach and response-adaptive randomisation informed by the posterior view of the dose-response profile.

## 2 Study structure

### 2.1 Intervention

The study intervention is the offer of a financial payment, delivered within a patient-support framework, for initiating DAA therapy for treatment of HCV. The payment will be made when the participant provides evidence of having initiated DAA therapy for HCV. The patient-support framework comprises Motivate-C study personnel (referred to as study navigators) that help patients navigate the process of initiating therapy. The offer is made at the time of randomisation (registration) and is paid on visual inspection by study staff of a prescription for DAA therapy within 12 weeks of registration. The amounts offered range from A$0 to 1000 in A$50 increments, assigned proportionally to the probability of effectiveness, see section Section 4.2. The range and increment were selected pragmatically on the basis that the largest amount is likely to be modest relative to the cost of DAA treatment and HCV management over the life of someone with HCV. Payment amounts are made via a digital debit card on the participants’ mobile device, or a physical debit card sent to the participants’ residence.

### 2.2 Endpoints

The primary endpoint is initiation of DAA therapy for the treatment of HCV infection within 12 weeks of registration, as evidenced by documented receipt of a valid dispensed DAA medication. Absence of such evidence at 12 weeks will be interpreted as a failure case, as will those participants that fail/decline to proceed further than registration.

The secondary endpoints are:

- Evidence of having had an HCV PCR test within 12 weeks of registration (regardless of result)
- Self-reported number of scheduled/recommended days of DAA not taken grouped into:
  - <7 days,
  - 7-13 days,
  - 14-20 days,
  - 21-27 days
  - 28+ days
  as reported within 14 days after the expected end date of DAA therapy (DAA therapy usually has a duration of 2-3 months)
- Sustained virological response (SVR), defined as a negative HCV RNA PCR test at any time from at least 4 weeks after completion of DAA therapy to before 12 months after registration.

### 2.3 Study population

The study population is defined in the protocol via the inclusion and exclusion criteria, summarised below. Inclusion criteria:

- 18 years or older
- current HCV infection (by self-report)
- Medicare-eligible
- Australia resident (i.e. currently living in Australia)

Exclusion criteria:

- currently receiving DAA therapy (or have at any time in the last 6 months)
- previously enrolled in the project
- unable or unwilling to provide informed consent
- unable or unwilling to complete follow-up
- unable to commence treatment within 12 weeks of randomisation due to any contra-indication (e.g., pregnancy, or breastfeeding)
- tested for HCV (RNA or serology) in the previous 4 weeks

People who self-report that they meet the eligibility criteria are registered as part of the study. However, to participate in the supported care and be eligible for any payments, the self-reported eligibility criteria must be verified by a study navigator.

### 2.4 Randomisation

Those who self-report as meeting the eligibility criteria via text message are registered into the study and are randomised to an incentive amount, which is retained in a REDCap database. Participant randomisation occurs via a custom randomisation module developed by the study team and embedded in the Motivate-C management system. The system is hosted by University of Sydney and has secure access, full audit trial and redundancy.

The registered participant is contacted to finalise the registration process by a study navigator. Once the registration processes are finalised, the participant can nominate a prescriber or is offered a default prescriber (one from a list of known prescribers). Nominated prescribers are randomised to either a A$0 or A$100 incentive. The prescriber randomisation uses a pre-generated 1:1 permuted block randomisation with block sizes of 2, 4, 6, 8 and 10. Default prescribers are provided with an administrative payment to compensate them for their involvement in the study.

The first enrolment cohort that register will be randomised with equal probability/weight to each incentive amount. However, at each interim analysis, the allocation weights will be updated based on a function of the modelled dose-response curve. The control arm (zero incentive amount) will be allocated a fixed weight, equal to the reciprocal of the number of unique incentive amounts. For all other arms, the randomisation weights will be calculated as described in Section Section 4.2. If an intervention’s posterior probability of effectiveness (defined as the probability of exceeding the minimum effective dose threshold value of 10%) falls below a pre-specified threshold, then the allocation for that intervention may be set to 0 and thus the intervention amount would effectively be dropped from the study. Consistency between the target weights and the desired assignment distribution is achieved via use of the mass-weighted urn design (Zhao 2015) and the allocation weight for the zero dose will remain fixed throughout.

Only specified analysts and a small number of administrative staff will be aware of the randomisation weights.

### 2.5 Sample size

As the intervention is the payment of a financial payment on evidence of initiating treatment, the sample size will be determined by the dose-response curve that is realised (capped by the budget received from the Medical Research Future Fund). For example, if the dose-response moves from its lowest to highest response at relatively low incentive amounts, then participants will be preferentially allocated to these lower amounts. Conversely, if, on average, higher incentive amounts are required to motivate treatment, then participants will be preferentially allocated to these higher amounts. Through simulation over a range of scenarios, we anticipate a total sample size of at least 1200 participants. This number of participants will achieve at least an 80% probability of being able to identify effective incentives (defined as an absolute increase of 10% in the treatment initiation rate relative to the zero-incentive response) when the true difference between the upper and lower bounds of the probability of response is 0.2 and a 6% type-I error rate for the null case, where no effective incentive amount exists.

### 2.6 Blinding

Given the nature of the intervention, blinding participants to their allocated treatment is not possible. However, the outcome can not be easily influenced by any stakeholder in that the primary endpoint is based on visual inspection of a standard medical prescription. Analysts and navigators are unblinded to the intervention due to specific tasks that they are required to perform.

Information on a participant’s allocated intervention is not provided to their nominated or default prescriber by the study team. This does not, however, prevent the participant from freely volunteering this information to their allocated prescriber.

### 2.7 Allocation concealment

As noted earlier, allocations are made by a centrally located randomisation module. The module is available only internally to privileged University staff and is called via the Motivate-C management software, deployed on secure University of Sydney servers.

## 3 Statistical considerations

### 3.1 Analysis populations

The primary analysis population will include all registrants (all registrants have been randomised) that have reached the primary endpoint. Participants providing evidence of DAA therapy initiation within 12 weeks of randomisation are success cases and all others are considered failure cases. This primary analysis population will be used for all outcomes, unless specified otherwise, and will follow the intention-to-treat (ITT) principle. All participants will be included and analysed according to the incentive they and their prescriber were initially allocated, irrespective of any deviations (e.g. ineligibility) from this or any other protocol deviations. However, the following exclusions will apply:

1. Participants who have been randomised but have not provided evidence of DAA therapy initiation and not yet reached 12 weeks post randomisation, will be excluded. These could be registrants that have not yet been contacted by study navigators and participants that have not provided evidence of treatment initiation for whom the 12-week timeline has not yet elapsed.
2. Any re-randomised participants (for any reason) will be excluded.
3. Participants who have met the primary endpoint, but for whom information has not yet been entered into the system will be treated as missing until the data has been entered.

### 3.2 Sequential analyses

The initial analysis will occur when the first 6-months of participants have reached the primary endpoint (12 weeks post registration). Based on an enrolment rate of 2 per day and a linear ramp up to this rate over the first 6-months, this equates to approximately 170-200 participants being observed at the first interim, distributed evenly over the available incentive amounts. If less than 100 participants are available for analysis at 6-months, then the first analysis will be deferred at monthly increments until a minimum of 100 participants have reached their primary endpoint.

Subsequent interim analyses are scheduled to occur every 4-months or after every A$100k committed to participant incentives, whichever occurs first. This may be varied as funds start to run low in order to ensure there are sufficient resources to provide incentives and administrative payments to both participants and clinicians. At each interim analysis there will be some participants who have registered but have not reached the required follow-up; these participants will not be included in the current interim analysis, but will be included in subsequent ones.

Only the primary analysis will be performed at each interim and the results will be used to compute the weights for the response-adaptive randomisation.

### 3.3 Descriptive summaries

The populations most impacted by hepatitis C are people who currently, or previously, inject drugs, prison populations, sex workers, people in rural and remote settings, and migrants from high-prevalence regions. These people constitute a marginalised group in society and are known to be wary of stigmatization. Therefore, the protocol has specified a minimal of set of data to be collected on participant characteristics. The purpose of this approach is to ensure minimal barriers to enrollment by limiting requests for personal details.

The only characteristics that will be collected from the participants include sex, age and state/territory of enrolment. Descriptive statistics on these variables will be calculated and presented by incentive amount, which may be aggregated for the sake of presentation. Categorical variables will be summarised by counts and proportions and continuous variables will be summarised either by minimum, maximum, means and standard deviations or medians and interquartile ranges. The number of missing values will be reported.

Additional descriptive statistics will be provided for:

- Primary and secondary outcomes.
- Number of participants registered over time (overall and by state) by incentive amount.
- Dollar amounts committed, issued and remaining.
- Number of participants who nominate prescriber versus accepting default prescriber

### 3.4 Participant flowchart

A participant flowchart will report on the following:

- registrations
- number of registrants contacted by navigator
- eligibility verified by study team navigator
- consented by study team navigator
- number of registrants proceeding to finalise enrolment
- number of participants accepting default versus nominated prescriber
- PCR status
- DAA status

### 3.5 Primary analysis

The primary analysis estimates a dose-response curve using a model-based approach. We assume the following specification:

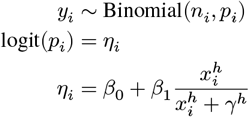

where

- *y*_*i*_ is the number of treatment initiations under dose *i*
- *n*_*i*_ are the number of participants allocated to dose *i*
- *p*_*i*_ is the probability of treatment initiation under dose *i*
- *η*_*i*_ is the log-odds of treatment initiation under dose *i*
- *β*_0_ is the log-odds of treatment initiation at the zero dose
- *β*_1_ is the difference between the lower and upper limits of response (in log odds)
- *x*_*i*_ is the incentive amount scaled to range between [0, 1] for dose *i h* is the hill parameter (controls the shape of dose repsonse)
- *γ* is the dose that produces a response halfway between the lower and upper limit

assuming the following priors

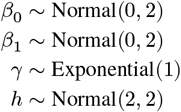

The equivalent Bernoulli parameterisation may be used.

It is possible that other initiatives designed to facilitate treatment for HCV will become active over the duration of the motivate C study. Given the use of response adaptive randomisation, this can lead to the temporal confounding (Lipsky and Greenland 2011). For example, assume that the intervention in Motivate-C leads to a small increase in the probability of treatment initiation and that this effect is observed at the first analysis. In response to this, the allocation weights would be updated, which leads to unbalanced allocation across the intervention arms in the next enrolment period. If an external initiative resulted in a change to the baseline probability of participants to initiate treatment over time, then this can bias the Motivate-C treatment effects when the data over the multiple enrolment cohorts are aggregated and analysed.

Adjustment for temporal variation is complicated in dose-response studies by virtue of the dependencies between the parameters in the model. Therefore, we do not adjust for temporal variation in the primary analysis, but we will include a sensitivity analysis that will characterise the time variation in the *β*_0_ and *β*_1_ terms. This analysis will assume independent first order random walks on these parameters, with appropriate scaling for the duration of time between interim analyses.

The construction of each of these terms is as follows: assume that *j* = 1, … *J* indexes each of the enrolment cohorts with *j* = 1 indexing the first cohort and *j* = *J* indexing the most recent cohort (the current cohort). For the current cohort, we make no adjustment to the *β*_0_ nor the *β*_1_ terms. That is, if *κ*_*m*,*J*_ denote the adjustments to *β*_0_ and *β*_1_ respectively, then we set *κ*_0,*J*_ = *κ*_1,*J*_ = 0. For the previous cohorts we ‘walk backwards’ computing 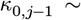 Normal 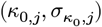 where 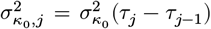 and 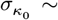 and where τ_*j*_ − τ_*j*−1_ represents the interval in time between the start of the *j*^*th*^ and the *j* − 1^*th*^ cohorts. We compute the *κ*_1,*j*_ using the same approach using an independent set of parameters.

We do not adjust for site as site is not applicable to the study.

#### 3.5.1 Effect of co-incentives

The effect of co-incentives for prescribers will be evaluated by accounting for group level heterogeneity in the *β* parameters from the base model. Specifically, three groups of participants are identified as those having received treatment from default prescribers, nominated prescribers (no incentive) and nominated prescribers (with incentive).

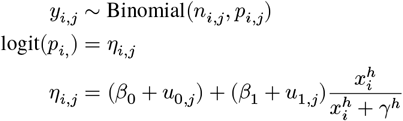

where the terms are as previously except now, we are considering each of the three groups *j* ∈ {1, 2, 3} and

- *u*_0,*j*_ is the group level offset from the overall lower limit of response term
- *u*_1,*j*_ is the group level offset from the overall difference between the lower and upper limits

with

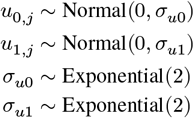

Other models may be fit and reported to supplement the primary analysis results, e.g. a multivariate perspective across all parameters or the introduction of a hierarchical model to enable deviations from monotonicity, see Gajewski et al. (2019).

### 3.6 Secondary analyses

#### 3.6.1 HCV PCR on bloods within 84 days after randomisation

Evidence of having had an HCV PCR test within 12 weeks of registration (regardless of result) will be represented as a binary indicator variable for each participant.

Given the manner in which the primary outcome is ascertained, this will be likely be approximately equal to the number of participants initiating treatment. For participants who seek treatment for their HCV, we could observe either positive or negative results. The denominator used will be the number of people randomised and we will simply report the data descriptively by incentive amount.

#### 3.6.2 Number of DAA days missed

Self-reported number of scheduled/recommended days of DAA missed within **14 days** of the recommended end date of therapy will be represented as an ordinal variable with the following levels

- less than 7 days
- 7-13 days
- 14-20 days
- 21-27 days
- 28+ days

as reported within 14 days after the expected end date of DAA therapy (DAA therapy usually has a duration of 2-3 months).

We will model this outcome using a cumulative logistic regression model with a covariate for the assigned participant incentive value and adjustment for co-incentive amounts offered to the prescriber. Specifically, we will adopt the stan parameterisation for the ordered logistic distribution.

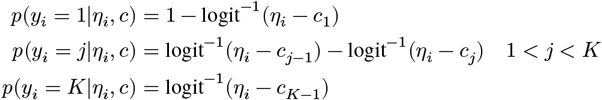

for categories *k* = 1..*K* and with *c*_*k*_ < *c*_*k*+1_ where *y*_*i*_ corresponds to the observed outcomes (category) for individual *i*. The individual level linear predictor will be constructed as

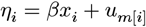

for participant *i* under incentive *x*_*i*_ and in group *m*[*i*] representing treatment under a default prescriber, nominated prescriber (no incentive) and nominated prescriber (with incentive) respectively. We will use regularising priors

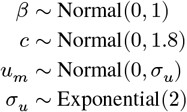

Sensitivity analyses may be explored such as using spline function rather than assuming linearity and excluding the group level heterogeneity components from the model.

#### 3.6.3 Sustained virological response (SVR)

SVR, defined as a negative HCV RNA PCR test at any time from at least 4 weeks after completion of DAA therapy and before 12 months post-randomisation will be represented as a binary indicator variable. Positive results and registrants failing to initiate DAA therapy or producing any evidence of an SVR result will be interpreted as failures. The outcome will be modelled using the same approach as the primary outcome. This analysis will be on all registrants that are randomised.

### 3.7 Economic analyses

In addition to characterising the dose response relationship between incentives and the probability of treatment initiation, the Motivate-C study will identify an optimal incentive via a cost-effectiveness analysis. The analysis will take a health services perspective with reference to a future cohort of participants. Costs will be expressed in Australian dollars in current prices at the time of the analysis and health outcomes will be expressed in terms of QALYs. Future costs and health outcomes will be discounted to the current value via a standard discounted cashflow perspective using a range of discount rates (3-7%).

The analysis will proceed by computing the total incentive and administrative costs for those that initiate treatment at each incentive amount. For those initiating treatment we will estimate the net present value of the lifetime management costs attributable to HCV based on their initial state along with the assumptions that DAA therapy is completed, and that they are cured. For those that do not initiate treatment, we will estimate the net present value of the lifetime management costs attributable to HCV, again based on the initial state, the assumption that DAA therapy is never initiated/completed, and that disease progression continues unabated.

The analyses are limited by the availability of participant-level data. Therefore, we adopt a proxy for this information from the current literature on the HCV population in Australia and model disease progression under a homogeneous Markov state transition model. Nominally, we will use the transition matrix based on Scott et al. (2022):

|  | f0 | f1 | f2 | f3 | f4 | dcc | hcc | transplant | death |
| --- | --- | --- | --- | --- | --- | --- | --- | --- | --- |
| f0 | 0.88 | 0.12 | 0.00 | 0.00 | 0.00 | 0.00 | 0.00 | 0.00 | 0.00 |
| f1 | 0.00 | 0.91 | 0.09 | 0.00 | 0.00 | 0.00 | 0.00 | 0.00 | 0.00 |
| f2 | 0.00 | 0.00 | 0.88 | 0.12 | 0.00 | 0.00 | 0.00 | 0.00 | 0.00 |
| f3 | 0.00 | 0.00 | 0.00 | 0.88 | 0.12 | 0.00 | 0.00 | 0.00 | 0.00 |
| f4 | 0.00 | 0.00 | 0.00 | 0.00 | 0.91 | 0.06 | 0.02 | 0.01 | 0.00 |
| dc | 0.00 | 0.00 | 0.00 | 0.00 | 0.00 | 0.78 | 0.07 | 0.01 | 0.14 |
| hcc | 0.00 | 0.00 | 0.00 | 0.00 | 0.00 | 0.00 | 0.38 | 0.02 | 0.60 |
| transplant | 0.00 | 0.00 | 0.00 | 0.00 | 0.00 | 0.00 | 0.00 | 0.50 | 0.50 |
| death | 0.00 | 0.00 | 0.00 | 0.00 | 0.00 | 0.00 | 0.00 | 0.00 | 1.00 |

which may be modified at the time of the analyses if there is are more recent data available that are relevant to the Australian population for HCV disease progression. Costs associated with each Metavir states will similarly be derived from Scott et al. (2022).

For health outcomes, we convert disease states to QALYs, again taking a lifetime view. Metavir state to QALYs will be derived from the Tufts Medical Centre Cost Effectiveness Analysis (CEA) registry.

Cost-effectiveness measures will be summarised as Incremental Cost-Effectiveness Ratios (ICERs) using those treatments that are neither strongly (those with higher costs and lower benefits than other treatment options) nor weakly (those with higher ICERs than more effective treatment options) dominated. The ICERs will be visualised as a cost-effectiveness frontier. From a cost-effectiveness perspective, the optimal treatment is that which has the highest cost-effectiveness relative to all other cost-effective treatment options. Equivalently, a decision-theoretic approach will be used to inform the problem of identifying the best treatment under uncertain information, see Baio (2013). For this we adopt the net (monetary) benefit as a utility function, defined as

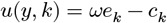

where

- *e*_*k*_ denotes the clinical effectiveness in terms of the present value of lifetime QALYs by dose incentive index *k*, drawn from a hypothetical future population
- *c*_*k*_ denotes the aggregated costs associated with the health outcomes
- *ω* denotes the decision makers willingness-to-pay

In this framework, the value of taking decision *k* is the expected utility and presupposing a risk neutral decision maker, the optimal incentive amount is that which maximises the expected utility subject to the decision makers willingness to pay. The multi-treatment equivalent of cost effectiveness acceptability curves will be prepared to characterise the uncertainty of a given treatment recommendation under imperfect information. The value of perfect information (VOPI) may be explored.

Sensitivity analyses will be undertaken to assess the impact of discount rates and unit-cost assumptions.

### 3.8 Safety analyses

The intervention is the commitment of a monetary amount on evidence of commencement of DAA therapy and is therefore considered to be of negligible risk to the participants and prescribers. However, an attempt to ascertain adverse events will occur when the navigator contacts the participant at 2-4 weeks after treatment initiation. Adverse events will be summarised descriptively only.

### 3.9 Model checking

Standard diagnostics will be run on MCMC chains for convergence. Models will be checked for goodness of fit. Model assessment and comparison will be via posterior predictive checks and cross-validation. Issues with convergence or stability may necessitate re-specifying or re-parameterising models. Additional models may be investigated as part of checks of sensitivity, stability, and model fit.

Any changes to the pre-specified models will be reported with a rationale for the changes that were made.

### 3.10 Missing values

Missing data can result from potential participants being randomised but not being contactable and also from loss to follow up at any point through the participant progression to the final SVR test. Within Motivate-C, missingness is generally interpreted as a failure event.

## 4 Statistical quantities and trial adaptations

### 4.1 Effective and futile doses

The effectiveness of an incentive amount is defined in terms of the posterior probability that the incentive increases the probability of response by more than 0.1 relative to the zero dose. An effective dose is one for which the posterior probability of effectiveness is greater than or equal to 0.95. We define the minimum effective dose as the lowest incentive amount that is deemed to be an effective dose.

Mathematically, the minimum effective dose k from the K design points is defined as min{*k*|(Pr(*p*_*k*_ − *p*1 + δ) > 0) ≥ ϕ}, where

- *p*_*k*_ represents the probability of treatment initiation under the incentive dose *k*
- *p*_1_ denotes the probability of treatment initiation under the first incentive amount, which is the zero-dollar dose
- δ equates to an increment of 0.1
- and ϕ is a threshold value for the posterior probability, which we have set at 0.95

A futile dose is defined as any incentive amount for which the probability of effectiveness is less than 0.2.

The only stopping rule relates to futility and exists to reduce the design space over time and to suspend enrolment if it becomes apparent that there is no substantive association between increasing financial incentives and the probability of treatment initiation.

Specifically, futility at the level of the study will be indicated if at any time from the third analysis, all incentive amounts are indicated as futile per the above definition.

### 4.2 Response adaptive randomisation

Initially, all participants are allocated with equal probability across the design space, which is restricted to the range A$0 to 1000 in A$50 increments. At the first interim analysis, we will update the allocation probabilities such that we will preferentially assign participants to incentives with a high probability of exceeding the minimum effective dose or better. The allocation probabilities for the prescriber incentives will remain fixed at 1:1 (incentive vs no incentive) for the duration of the trial.

The participants dose allocation algorithm computes the weights proportional to the product of the probability of effectiveness for each dose and the first differences between responses (on the probability scale) for successive doses.

The weights will be set to zero if dose response is found to be declining. Such a scenario would likely necessitate a variation to the study design.

The calculation uses the standard variance weighted approach:

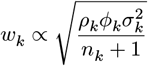

where

- *ρ*_*k*_ corresponds to the probability of effectiveness (i.e. ≥ MED threshold)
- ϕ_*k*_ corresponds to the first difference, namely ϕ_*k*_ = *p*_*k*_ − *pk*−1
- *σ*_*k*_ is the standard deviation of the posterior log-odds of response and
- *n*_*k*_ is the number of enrolments reaching the primary endpoint for arm *k* = 2 … *K* − 1.

These weights are transformed to sum to 1 − (2/*K*) and give the allocation probabilities with the zero dose and maximal dose allocation remaining fixed at 1/*K* throughout the entire study.

## 5 Data Sources

Data collection is partially automated (via SMS interactions) and partially entered by study personnel based on participant self-report. Data are stored in a Motivate-C REDCap database and some (non-identifiable) auxiliary data are retained in separate proprietary database. Data necessary for the analyses and randomisation updates are stored within REDCap and will be provided as extracts or via a REDCap API token.

A data management plan has been developed and the Trial Master File is held centrally.

## 6 Quality control

Quality control will primarily comprise code reviews as well as review of the analysis reports by a second trial statistician and independent statistical monitor.

Unit tests have been produced for the randomisation module and will be updated over time as required. Informal integration testing with the other system components will be undertaken in a development environment.

## 7 Reporting

The final results from the project will be communicated by presentations (including seminars and conference presentations) and publications.

Interim analysis results will be documented in a report format, including a summary of the current study status, enrolment progression, descriptive summaries, the results from the primary analysis and the revised allocation probabilities. These reports will quality checked by the second trial statistician and forwarded to a statistical monitor for review and their recommendations.

The safety extract and summary, which is unblinded to incentive amounts, will provided to the independent safety monitor.

## 8 Declarations

### 8.1 Trial registration

Motivate-C was registered at ANZCTR (anzctr.org.au), Identifier ACTRN12623000024640 on 11 January 2023 (https://anzctr.org.au/Trial/Registration/TrialReview.aspx?id=384923&isReview=true).

### 8.2 Author contributions

All authors involved in concept and design. MJ lead design specification and documentation. All authors reviewed, provided feedback and approved the manuscript for submission.

### 8.3 Author approval

All authors have reviewed and approve this statistical analysis plan in its present form.

## 8.4 Acknowledgements

The authors acknowledges Dr Liz Ryan and Dr Robert Mahar for their contributions as independent statistical monitors and critical review of the design specification. The authors also acknowledge the broader Motivate-C team who have been involved in discussions regarding the development of study.

## 8.5 Ethics

This study was approved by the Sydney Local Health District Human Research Ethics committee (Ref: 2022/ETH0168).

## 8.8 Data Availability Statement

There are no data associated with the preparation of the analysis plan. De-identified participant level data arising from the study can be made available after study completion with appropriate ethics approvals.

